# An epidemic model for economical impact predicting and spatiotemporal spreading of COVID-19

**DOI:** 10.1101/2020.09.02.20186551

**Authors:** Mateo Cámara, Mario Miravete, Eduardo Navarro

## Abstract

Since the emergence of a new strain of coronavirus known as SARS-CoV-2, many countries around the world have reported cases of COVID-19 disease caused by this virus. Numerous people’s lives have been affected both from a health and an economic point of view. The long tradition of using mathematical models to generate insights about the transmission of a disease, as well as new computer techniques such as Artificial Intelligence, have opened the door to diverse investigations providing relevant information about the evolution of COVID-19. In this research, we seek to advance the existing epidemiological models based on microscopic Markov chains to predict the impact of the pandemic at medical and economic levels. For this purpose, we have made use of the Spanish population movements based on mobile-phone geographically-located information to determine its economic activity using Artificial Intelligence techniques and have developed a novel advanced epidemiological model that combines this information with medical data. With this tool, scenarios can be released with which to determine which restriction policies are optimal and when they have to be applied both to limit the destruction of the economy and to avoid the feared possible upsurge of the disease.

## Introduction

As of 22 January 2020, the outbreak of coronavirus disease has infected more than 6.400.000 people worldwide, raising more than 370.000 deceases [1]. Epidemiological models are a mathematical approach used to analyze the evolution of infectious viral processes and to predict the development of these in the future [2]. They are based on the usage of statistics, assumptions and data to adjust mathematical parameters so they suit the most to the epidemic or the particular phase of a disease. It is an integral part of the preliminary studies carried out by competent bodies to understand the soundness of possible interventions that may be carried out, both from a social and an economic point of view [3,4]. It is precisely this point one of the biggest problems to address since it has a direct impact on many people’s lives. Making intelligent use of data optimizes decisions regarding the control of the epidemic.

Epidemiological models have been developed from the available data both to understand the disease and to predict what would happen in the future [5]. One of the classic models used as a basis for many more advanced ones is the SIR model [6,7]. The acronym refers to “susceptible”, “infected” and “recovered”, which are the possible states in which a subject can be found within a specific population. Based on each of these conditions, a series of differential equations are drawn to define how a subject can step forward from one state to another. The basic SIR model assumes that the population is randomly mixed within itself, dealing with early assumptions that usually never happen in the real world. Since it is not a very realistic human behavior, some more complex ones have been defined around it, such as the MSIR model –which includes immunity to birth- [8], SEIR –including exposed state- [9] or SIX – in which there is no immunity [10]. SIR principles set the basis for most of the epidemics state of the art. However, it is important to notice that many other dimensions may lead to human interactions, rather than just geographical locations. Populations can be structured into age, risk or ethnic groups. Even though some authors have modeled these facts as a geographic dimension [11,12], a whole scientific branch has been developed out of it, reaching what it is known as metapopulation spatial models [13–16]. These are represented as networks with groups considered as nodes and interactions modeled like links. This general approach to the state of the art settles the basis of the presented research, which will make use of a Microscopic Markov Chain approach to handle both metapopulation dimensions and possible disease states.

In the specific case of the epidemic due to COVID-19 disease, numerous recent studies have sought to model the evolution of the disease by stratifying the population according to its demographic and spatial distribution, age and previous pathology [17–19]. They present each of these parameters within a Microscopic Markov Chain approach formulation with specific states to identify the spread of the virus in complex human networks [20,21]. Some other interesting studies rely on Artificial Intelligence techniques to forecast the contagion trajectory of COVID-19 [22–24]. These recent studies have served as a basis for our research. In particular, the paper presented by the University of Zaragoza [25] has been the scientific basis on which we have tried to push forward. Our research is novel to the extent that we have sought to advance, optimize and improve as much as possible the principles on which their research is focused.

It has been shown that the particularities of COVID-19 have forced a sharp reaction and an intense search for the causes of its rapid expansion throughout the world [26–28]. The effect of asymptomatic people has been considered a fundamental vector of the virus that has led many infected people to continue living a normal life while keeping infecting others [29,30]. Cases of asymptomatic often occur in young people, who unknowingly bring the virus into the homes of their relatives with adult or elderly people who end up suffering the consequences. It also seems to be true that COVID-19 is transmitted through the air [33,34], so measures taken by governments and prevention institutions, such as the distribution of masks, disinfection of public spaces, or the prohibition of leaving homes, are considered critical [28]. Finally, people who are severely affected by the disease and require medical services are usually affected in their lungs, preventing their proper functioning and leading to episodes of drowning [35]. The use of respirators in Intensive Care Units has been essential to saving lives [36]. However, some countries have experienced high-risk circumstances when medical services collapsed [37,38]. People requiring them could be left unattended due to high occupancy. It is these three reasons that led to the development of an epidemiological model such as the one proposed in this publication: A model capable of taking into account the impact of asymptomatic people, air transmission dependent on population movements and the state of medical services. Besides, we have sought to make our system qualified for launching multiple possible future scenarios. For this purpose, it has been designed a complex set-up for adjusting parameters to the historical evolution of the COVID-19 employing a genetic algorithm based on Artificial Intelligence [39]. It also has been developed an interactive platform that allows users to manually indicate the possible value of the parameters in the future so that an expert team can produce reports based on this tool. It can be also used to monitoring and assessing the impact of confinement measures through contextualized mobility information. The underlying intention is to support the decision-making of health authorities for the epidemiological control to face large-scale biological emergencies and to discern the economical costs of it. In this regard, we have developed a Markov-based algorithm to automatically predict the percentage of the population that stands in each state of the disease for each defined region at each time instant. It has been tested and effectively applied to the Spanish population from March to October 2020.

## Materials and methods

In this section, each of the components that have constituted the designed system is examined. There are several differentiated parts. However, only their intended use will be mentioned except for the epidemiological model, which will be explained as detailed and precise as possible. Fig. 1 shows each of the blocks that make up the model and their connections. The system is composed of four large subgroups.

**Figure 1.**
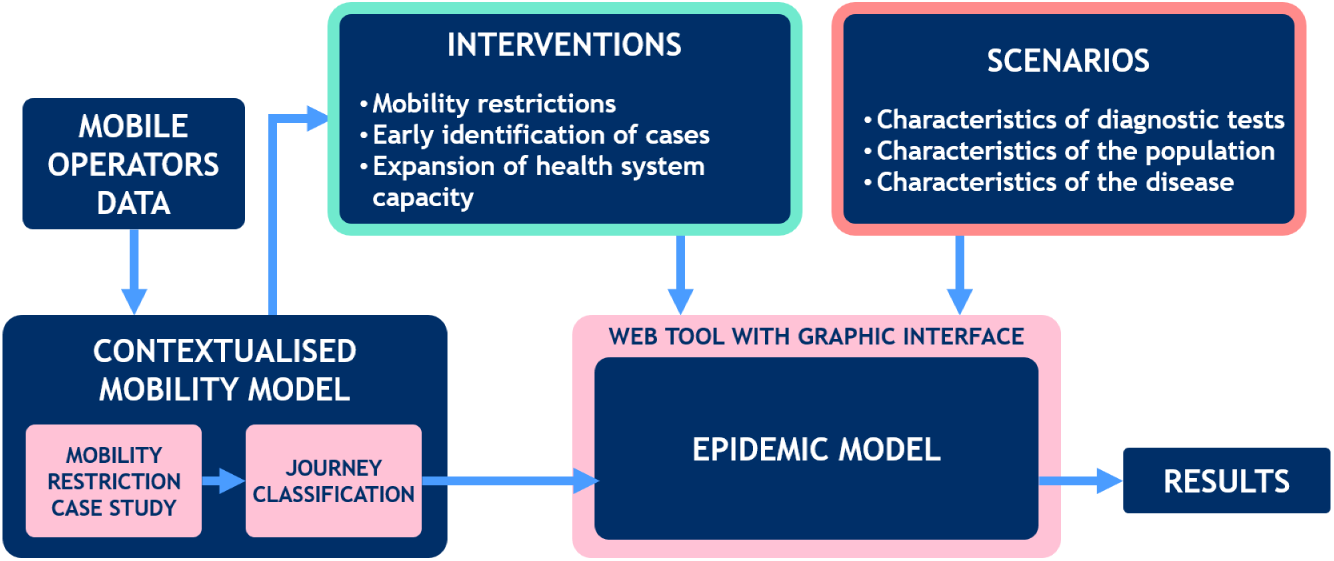
high-level block diagram. The epidemiological model forms the central core between the population mobility model, government interventions and possible disease evolution scenarios, leading to predictive results for the future.

- A contextualized mobility model. This is a series of unsupervised Artificial Intelligence algorithms that are capable of discovering typologies of people’s movements. People’s mobility data are partially tagged, making it easier to find patterns and associate them with a specific economic activity. They are obtained from anonymous mobile operators’ data that have been provided by the Spanish National Statistics Institute (INE) [40]. At the time the experiments were carried out, mobility data was available until May 31, 2020.
- An interactive model of social and economic interventions. This tool allows end-users to simulate all kinds of scenarios by modifying relevant parameters that may affect the spread of the epidemic such as control measurements.
- A scenario launcher. The proposed system is capable of calculating the parameters of the epidemiological model that best fit the historical evolution of the disease as a result of a genetic algorithm. However, it is also possible to manually assign certain parameters in order to understand possible new scenarios in the evolution of the disease. It is designed so that experts in the field can interact with different alternatives and proposals.
- An epidemiological model. Which will be detailed in-depth in the following subsection and which forms the core of this publication.

## Epidemiological model

We propose an epidemiological model that has been tested for COVID-19. Fig. 2 shows an outline of the model. It is based on the following concepts which form the initial assumptions of the system:

**Figure 2.**
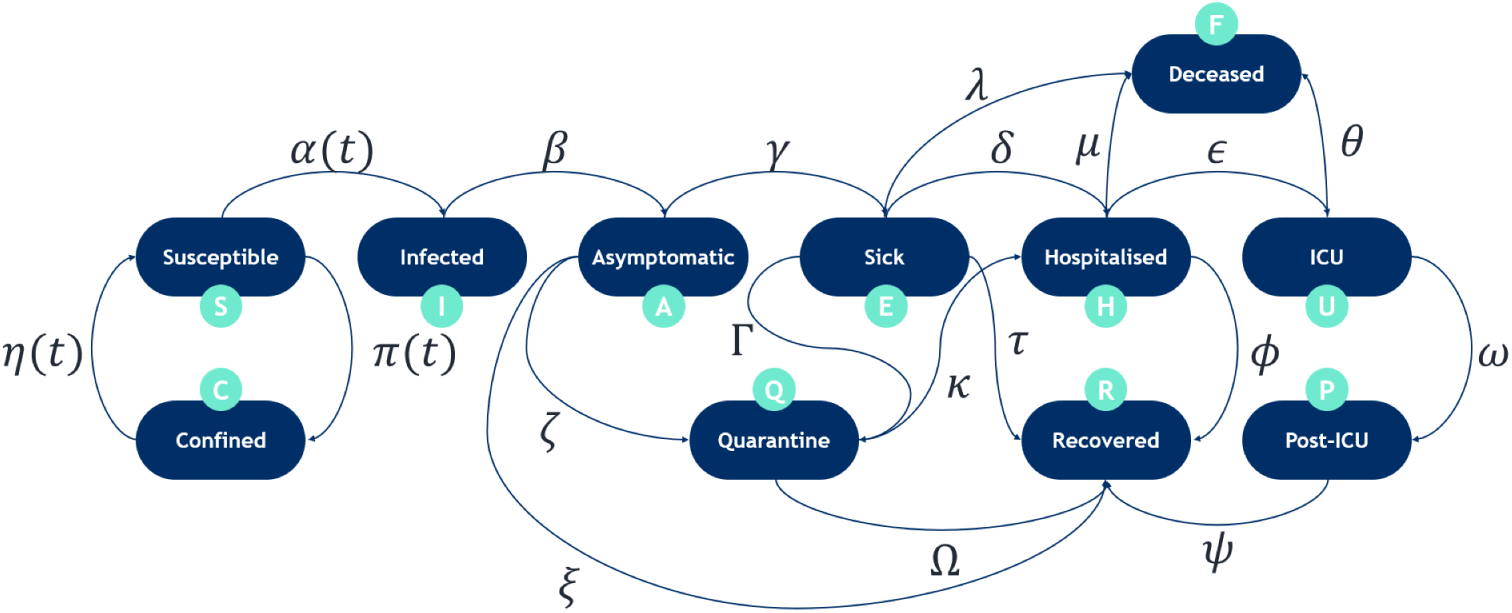
Epidemiological model. The epidemiological model forms the central core between the population mobility model, government interventions and possible disease evolution scenarios. It is composed of a total of eleven states.

- The population is divided into three age segments. It has been shown that the evolution of patients with COVID-19 is dependent on age range [1,32]. Infected young people are usually asymptomatic or have very mild symptoms that can be mistaken for the common flu [31]. Older age groups have a risk of more severe symptoms that are more likely to require the limited health services, which in case of collapse, could lead to the death of people who could not be assisted.
- The population is divided into three thousand geographical patches (in the case of Spain). Population movement has proven to be a determining factor since contagion occurs from person to person. The number of regions is determined by the dataset to be used. The higher the granularity, the better the accuracy in determining population movements. There is a compromise between the size of the region and the ability to anonymize the dataset, so it is common to find large sections. For example, a territory like Spain, counting more than 500 thousand square kilometers with a population of almost 50 million people is divided into only 3000 regions to guarantee the privacy of the population’s movements.
- Every person in the population must be located in one and only one state at a certain time. The epidemiological model counts with eleven states that represent each of the possible phases in which a person might be found. It considers that every person is susceptible to contracting the disease (no birth immunity).
- Each of the states is connected to at least other state.
- Connections between states are modeled as a time-fixed ratio except for those that connect Confined, Susceptible and Infected states.

The model incorporates states that allow the evaluation of the evolution of the disease in a context where the representation of asymptomatic patients has been very relevant. Besides, some hospital states are included to assess the saturation of medical services, both emergency and ordinary. Each of the states has a discrete-time dependency. Each time step corresponds to twenty-four hours.

The model contains eleven states that are defined as:

- SUSCEPTIBLE (S): a person who maintains his/her professional or academic activity and who has possibility to interact outside their region of belonging.

– Initial status of all persons not immune to COVID-19.
– Reached from confined state (C) at a rate *η*(t).
– Eq. (1) defines the total population in state S and handles the transition from one instant of time (defined by t) to the next for each age stratum (defined by g) and for each geographical patch (defined by i).

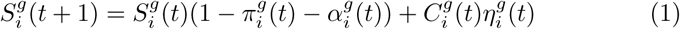
- CONFINED (C): a person who has suspended his/her professional or academic activity and who does not has the possibility to interact outside its region of belonging.

– Reached from susceptible state (S) at a rate *π*(t).
– Eq. (2) defines the total population in state C and handles the transition from one instant of time (defined by t) to the next for each age stratum (defined by g) and for each geographical patch (defined by i).

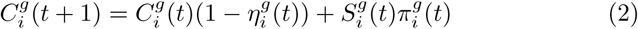
- INFECTED (I): an exposed person who is a carrier of the virus, but it’s not contagious yet.

– Reached from susceptible state (S) at a rate *α*(t).
– Eq. (3) defines the total population in state I and handles the transition from one instant of time (defined by t) to the next for each age stratum (defined by g) and for each geographical patch (defined by i).

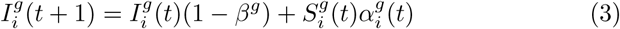
- ASYMPTOMATIC (A): a person who is a carrier of the virus, can transmit it and has mild symptoms (or no symptoms) of COVID-19 and carries on his/her usual activities.

– Reached from infected state (I) at a rate *β*.
– Eq. (4) defines the total population in state A and handles the transition from one instant of time (defined by t) to the next for each age stratum (defined by g) and for each geographical patch (defined by i).

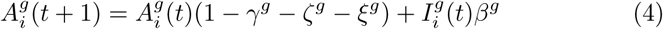
- SICK (E): a person who is a carrier of the virus and has symptoms of COVID-19. This status is not a transmission vector of the virus, since the person is isolated at home.

– Reached from asymptomatic contagious state (A) at a rate *γ*.
– Eq. (5) defines the total population in state E and handles the transition from one instant of time (defined by t) to the next for each age stratum (defined by g) and for each geographical patch (defined by i).

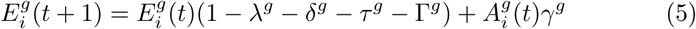
- QUARANTINE (Q): a person forced into confinement because they have been positively detected by COVID-19. Therefore, this state is not a transmission vector of the virus.

– Reached from:

* Asymptomatic state (A) at a rate *ζ*.
* Sick state (E) at a rate *ρ*.
– Eq. (6) defines the total population in state Q and handles the transition from one instant of time (defined by t) to the next for each age stratum (defined by g) and for each geographical patch (defined by i).

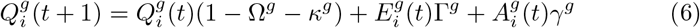
- HOSPITALIZED (H): a person using general hospital services (not specific emergency services).

– Reached from:

* Sick state (E) at a rate *δ*.
* Quarantine state (Q) at a rate *κ*.
– Eq. (7) defines the total population in state H and handles the transition from one instant of time (defined by t) to the next for each age stratum (defined by g) and for each geographical patch (defined by i).

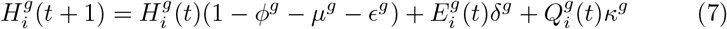
- ICU (U): a person using emergency hospital services of the Intensive Care Unit.

– Reached from the hospitalized state (H) at a rate *ε*.
– Eq. (8) defines the total population in state U and handles the transition from one instant of time (defined by t) to the next for each age stratum (defined by g) and for each geographical patch (defined by i).

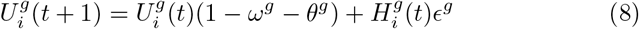
- POST-ICU (P): a person making use of general hospital services spending time while recovering from ICU services.

– Reached from ICU state (U) at a rate *ω*.
– Eq. (9) defines the total population in state P and handles the transition from one instant of time (defined by t) to the next for each age stratum (defined by g) and for each geographical patch (defined by i).

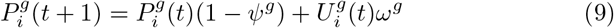
- RECOVERED (R): a person immune to COVID-19 after having overcome the illness.

– Possible final status of all persons who have been infected with COVID-19.
– Reached from:

* Post ICU state (P) at a rate *ψ*.
* Quarantine state (Q) at a rate Ω.
* Asymptomatic state (A) at a rate *ξ*.
* Sick state (E) at a rate *τ*.
* Hospitalized state (H) at a rate Φ.
– Eq. (10) defines the total population in state R and handles the transition from one instant of time (defined by t) to the next for each age stratum (defined by g) and for each geographical patch (defined by i).

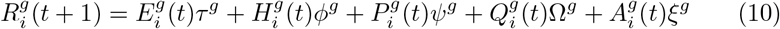
- Deceased (F): a person who died from COVID-19 causes.

– Possible final status of all persons who have been infected with COVID-19.
– Reached from:

* Sick state (E) at a rate of *λ*.
* Hospitalized state (H) at a rate *µ*.
* ICU state (U) at a rate *θ*.
– Eq. (11) defines the total population in state F and handles the transition from one instant of time (defined by t) to the next for each age stratum (defined by g) and for each geographical patch (defined by i).

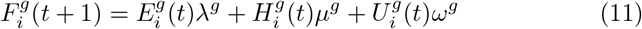

The rate 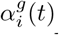 with which the population of the region *i* and the age group *g* transitions from the state susceptible (S) to non-contagious infection (I) is defined in Eq. (12). The value 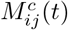 represents the mobility matrices for each of the c economical activities detected in a cluster formed by up to W different categories. Since governments are able to apply restrictions to certain economic activities represented by a cluster, there is a correction factor *b*^*c*^(*t*) with a value between 1 and 0 that represents to what extent activity c is allowed. When all matrices are sum up, a global movement matrix is obtained, assuming each person belongs to one cluster only for each instant of time. Eq. (12) is also affected by the probability that the agents get infected by the pathogen according to its geographical patch i and age group g, represented as 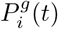.

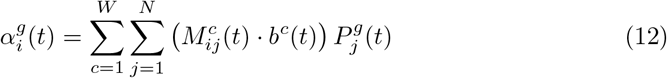

The probability of being infected in the region i, represented as 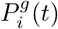, is calculated through the number of people in the Asymptomatic state *A*(*t*). It is defined in Eq. (13).

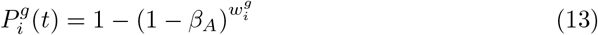

where 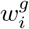 is the average number of contacts made by a person with asymptomatic people during one day; defined in Eq. (14).

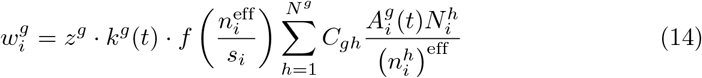

where 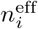 is the effective population in the age group *g* and patch *i, C*_*gh*_ is the contact matrix that captures the average percentage of contacts between age groups as defined in [25], *k*^*g*^(*t*) is the average number contacts by age, 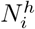 is the total population of the age group *g* and patch *i, s* is the area of the patch *i*, the function *f*(*·*) captures the influence of the population density and *z* is a normalization factor calculated as in Eq. (15).

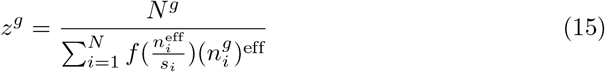

where *N*^*g*^ is the population of the age group *g* and 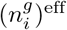 and 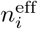 are defined, respectively in Eq. (16) and Eq. (17)

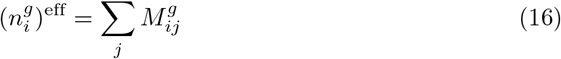

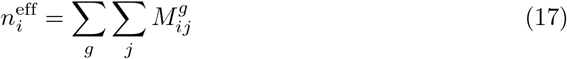

The effect of the population density is captured by [25,41] as in Eq. (18).

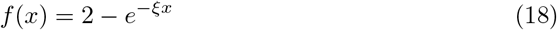

To assess the confinement of the population and the safe-distance restrictions at an instant *t* the following guidelines have been followed. First, the mobility matrix **M**^*g*^(*t*) is modified to adequate the number of people moving between regions within a day with the specific restriction policies at that moment. Second, part of the population in Susceptible state (S) is transferred to Confined state (C) and vice versa depending on the increasing or decreasing movement restrictions. Finally, the average number of contacts *k*^*g*^(*t*) is also modified. This setup accurately models most early situations in European countries such as Spain or Italy, where they have experienced hard lock-downs. Let us define *p*(*t*) as the mobility restriction parameter, therefore the average number of contacts, *k*^*g*^(*t*) and the mobility matrix are affected as in Eq. (19) and Eq. (20).

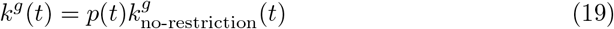

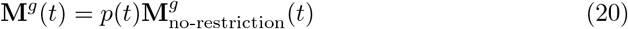

## Results & Discussion

In this section, we present the results of having applied our epidemiological model to the Spanish population taking as the starting date the first outbreaks of COVID-19 on February 20, 2020. In this paper, we present the results of two different scenarios in order to compare the importance of the restriction policies. Table 1 shows the parameters used in the epidemiological model for both scenarios. Each of them has been obtained through a genetic algorithm based on Artificial Intelligence (except some parameters which have been extracted directly from the state of the art), which has explored multiple options until reaching an adjustment to the historical data with a minimum mean square error. However, the ranges of values among which the algorithm could vary have been defined by hand within the realistic possibilities of each of the parameters, using the daily reports of the Spanish Ministry of Health [42]. We have considered that the disruption of COVID-19 in Spain has had a direct relationship with the number of asymptomatic infected people, as expressed in one of the lines of research that best fits the reality [43,44]. In our case, we have decided to simplify the model to the point that it only considers infections due to contact with asymptomatic people. Reality is not so far from this assumption since patients who present symptoms tend to self-reclusion in their homes and their infectivity is irrelevant to the analysis of society as a whole. On the other hand, there is no database of the mobility of people when their life has suffered an eventuality such as contracting an illness. Historical information up to 14 May 2020, was used as a training dataset to adjust the epidemic model to real values. Historical information up to 31 May 2020, was used as a validation dataset to assess how accurate the model was. Predictions have been computed until 26 October 2020 to understand to what extent there may be an upsurge of COVID-19 infections in Spain.

**Table 1.** Parameters of the model. All of them have been computed based on an iterative Artificial Intelligence algorithm except for those marked with a “*” since they have been extracted from several recent works.

| Age group | 0-14 | 15-64 | 65-120 |
| --- | --- | --- | --- |
| Percentage of transition from Asymptomatic to Sick | 30% | 75% | 75% |
| Percentage of transition from Asymptomatic to Recovered | 70% | 25% | 25% |
| Transition time from Asymptomatic to Sick | 4.5 | 4.5 | 4.5 |
| Transition time from Asymptomatic to Recovered infection | 8.5 | 8.5 | 8.5 |
| Percentage of transition from Sick to Deceased | 0.0% | 0.5% | 3.0% |
| Percentage of transition from Sick to Hospitalized | 2.0% | 3.6% | 11.7% |
| Percentage of transition from Sick to Recovered | 98.0% | 95.9% | 85.3% |
| Transition time from Sick to Deceased | 11 | 11 | 11 |
| Transition time from Sick to Hospitalized | 6 | 6 | 6 |
| Transition time from Sick to Recovered | 11 | 11 | 11 |
| Percentage of transition from Quarantine to Sick | 30% | 75% | 75% |
| Percentage of transition from Quarantine to Recovered | 70% | 25% | 25% |
| Transition time from Quarantine to Sick | 4.5 | 4.5 | 4.5 |
| Transition time from Quarantine to Recovered | 8.5 | 8.5 | 8.5 |
| Percentage of transition from Hospitalised to Deceased | 0.0% | 3.0% | 15.0% |
| Percentage of transition from Hospitalised to Recovered | 99% | 95% | 82% |
| Percentage of transition from Hospitalised to ICU | 1.0% | 2.0% | 3.5% |
| Transition time from Hospitalised to Deceased | 5 | 5 | 5 |
| Transition time from Hospitalised to Recovered | 10 | 10 | 10 |
| Transition time from Hospitalised to ICU | 2 | 2 | 2 |
| Percentage of transition from ICU to Deceased | 2% | 20% | 30% |
| Percentage of transition from ICU to post-ICU | 98% | 80% | 70% |
| Transition time from ICU to Deceased | 3 | 3 | 3 |
| Transition time from ICU to post-ICU* [45] | 10 | 10 | 10 |
| Transition time from post-ICU to Recovered | 5 | 5 | 5 |
| Infectivity of asymptomatics | 3.75% | 3.75% | 3.75% |
| $k_g$ - Average number of contacts by age* [40] | 11.8 | 13.3 | 6.6 |

In Spain, the ministerial authorities have provided information on the evolution of the coronavirus at Autonomous Community level. Spain is divided into a total of 19 Autonomous Communities in which the evolution of the disease has been different. They began to officially count the cases as of February 29. The number of detected cases was introduced in our epidemiological model, giving results incompatible with the reality of the following months. COVID-19 report from the Department of Epidemiology of the Imperial College in London [46] was taken into consideration, which determined that on a global level, the number of undetected cases could be up to 2/3 higher. Multiplying this rate by the official values we obtained the result shown in Table 2, which allowed a realistic adjustment of the epidemiological model.

**Table 2.** Initial asymptomatic people in each autonomous region. The number of asymptomatic people has been obtained as a function of the first data registered by the Spanish Ministry of Health.

| Autonomous region | Asymptomatic |
| --- | --- |
| Andalucía | 107 |
| Aragón | 65 |
| Asturias, Principado de | 33 |
| Balears, Illes | 23 |
| Canarias | 13 |
| Cantabria | 16 |
| Castilla y León | 230 |
| Castilla - La Mancha | 590 |
| Cataluña | 450 |
| Comunitat Valenciana | 78 |
| Extremadura | 130 |
| Galicia | 50 |
| Madrid, Comunidad de | 830 |
| Murcia, Región de | 12 |
| Navarra, Comunidad Foral de | 49 |
| País Vasco | 83 |
| Rioja, La | 20 |
| Ceuta | 1 |
| Melilla | 2 |

The scenarios presented in this document use the same epidemiological parameters shown in Table 1. The difference lies in the restriction of mobility, in the advice given to citizens and in the individual responsibility of people at the different stages of the epidemic. Our epidemiological model considers the movement of asymptomatic people as one of the main vectors of the disease. However, movement can be less dangerous if several circumstances relax the risk, such as the use of masks [47]. In Table 3 we show the values used in each of the scenarios as a percentage in which mobility is maintained. However, people who move safely (maintain a safe distance and/or wear a mask) are considered to be not in danger of contagion. Therefore, the percentages affected both mobility and average contacts per person *k*^*g*^. The differences between the two scenarios appear from May onwards. In scenario 1, the Spanish population regains the right to move freely within the national territory, however, it is not aware of the possibility of contagion. So from 21 June, the day on which Spain revokes its state of emergency, the population practically lives a normal life similar to that before the COVID-19. In scenario 2, the population maintains a very high level of hygiene and awareness, and although it is legal to move freely from 21 June, they do so with moderation and conscience. The government has a lot to contribute to these policies, as it can encourage this awareness and promote the use of masks. We are aware that these events are impossible to measure, but we find the results of the underlying idea very interesting.

Figs. 3 and 4 show the aggregated quantitative results at the national level for the two different scenarios. In the first scenario, it can be seen that the number of cases is rising again because the population has not taken the necessary preventive measures and has caused a new peak of contagion. In the second scenario, people who were infected were aware of the risk and although the dangerous mobility is up to 30%, there is no upsurge, probably leading to the disappearance of the virus after some time. Particularly noteworthy is that the absolute number of cases in the resurgence scenario reaches approximately the same severity as the first peak. However, no preventive measures have been simulated at the time of the outbreak, and yet the peak is reduced. This interesting result may be because that cluster immunity has been achieved. Nevertheless, the cost in human lives is very high, doubling the number of deaths.

**Table 3.** Mobility restrictions. These values depend on the measures taken by the Spanish government, from the moment the alarm state started, until the end of the forecast.

| Starting day | Scenario2 | Scenario 1 |
| --- | --- | --- |
| 20-Feb | 100% | 100% |
| 10-Mar | 75% | 75% |
| 14-Mar | 32% | 32% |
| 28-Mar | 28% | 28% |
| 12-Apr | 30% | 30% |
| 07-May | 50% | 30% |
| 14-May | 50% | 30% |
| 21-Jun | 75% | 30% |

## Conclusion

In this paper, we have presented an epidemiological model based on Microscopic Markov chains that are fed with geolocalized population information and demographic data. It is a tool that has emerged from the irruption of the COVID-19 in the world and that aims to serve as an extra element for prediction and decision making by competent bodies. Our system has the capacity to determine the economic activities of the population based on their movements, so intelligent containment measures can be applied.

It can be seen how the influence on the age of the subjects is very relevant. People in the 65–120 age group often need medical services, while those infected at other ages tend to recover without using them. The infectivity of asymptomatic people is an issue that is still not sufficiently known. Our algorithm has determined that each person infects an average of 3.75% of the people they contact. This number is close to the proposed values in the state of the art which vary from 3% to even 10%.

The measures adopted by private companies, which favored teleworking as much as possible, have also been taken into account, as well as the measures of some regional governments to provide the population with masks, which reduce the infectivity of sick people and which have been modeled as a reduction in their movement. We consider these measures to be very favorable to people’s health. They will help to ensure that the epidemic does not spread easily and that the economy does not suffer such devastating effects.

Given the results, it seems clear that the Spanish population is at risk of suffering a resurgence at the end of the summer. Hygiene measures and social distance should be a priority for all people. Strict confinement is a possibility until a vaccine is found, except that it damages the economy in a very harmful way, and more so in the case of Spain whose GDP depends largely on tourism in summer. Therefore, reaching a compromise should be the priority of the competent authorities. Herd immunity is still far from being a reality, so it is unfeasible to lead a normal life without expecting a new collapse of health services. Giving clear information to the population about the risks of coronavirus is a determining factor in avoiding many infections, as well as providing masks and disinfecting meeting spaces. The individual responsibility of an informed society may be sufficient to achieve herd immunity gently, or in its absence, to achieve vaccination, in both cases without collapsing health systems or destroying the economy.

**Figure 3. Scenario 1.**
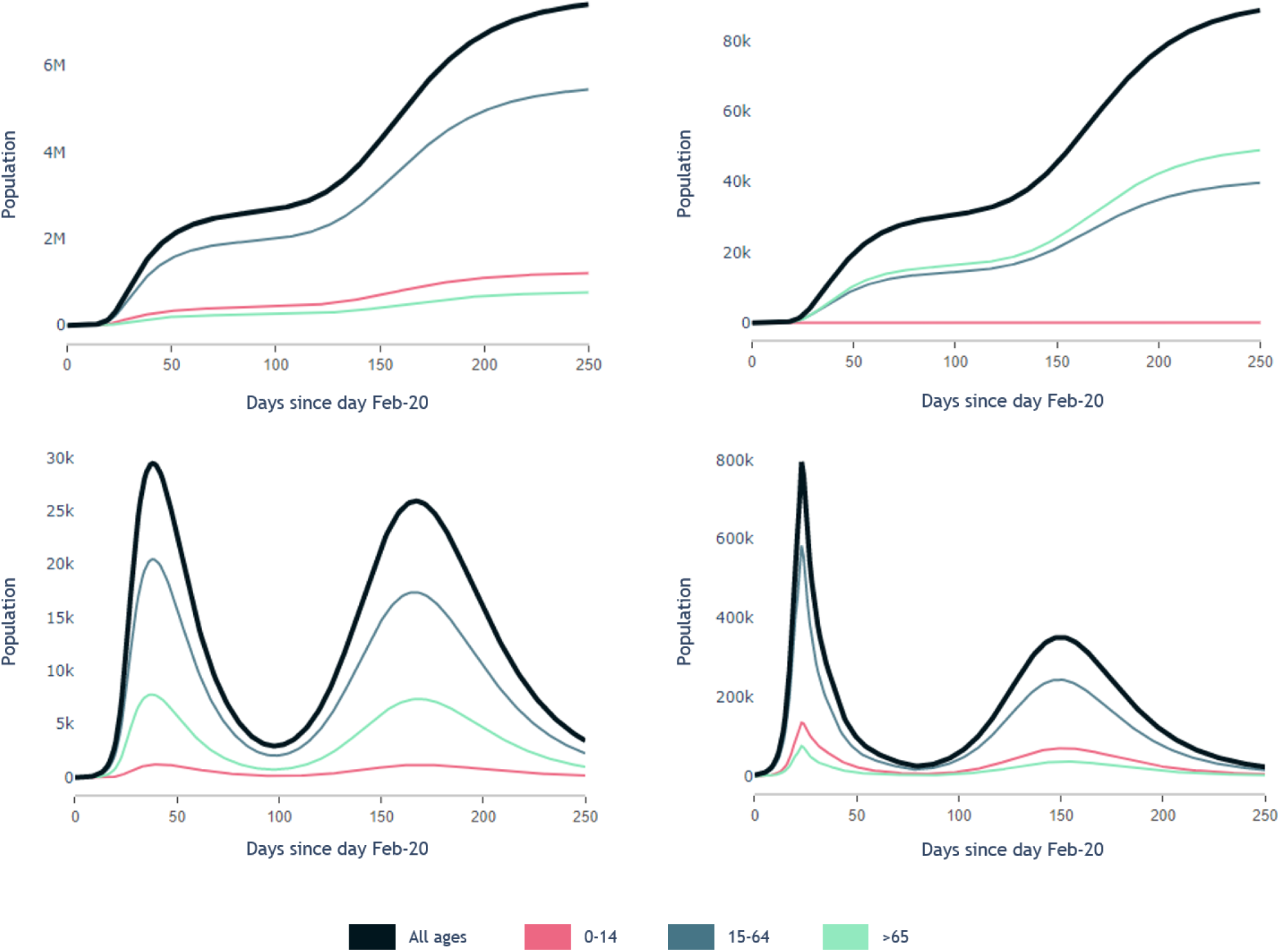
Mobility restricted up to 30% of movements and later released up to 75%. From left to right, up to down, first graph shows accumulated number of people who recovered after contracting COVID-19, second graph shows accumulated number of people who deceased by COVID-19, third graph shows instantaneous number of people making use of health services and fourth graph shows instantaneous asymptomatic people.

**Figure 4. Scenario 2.**
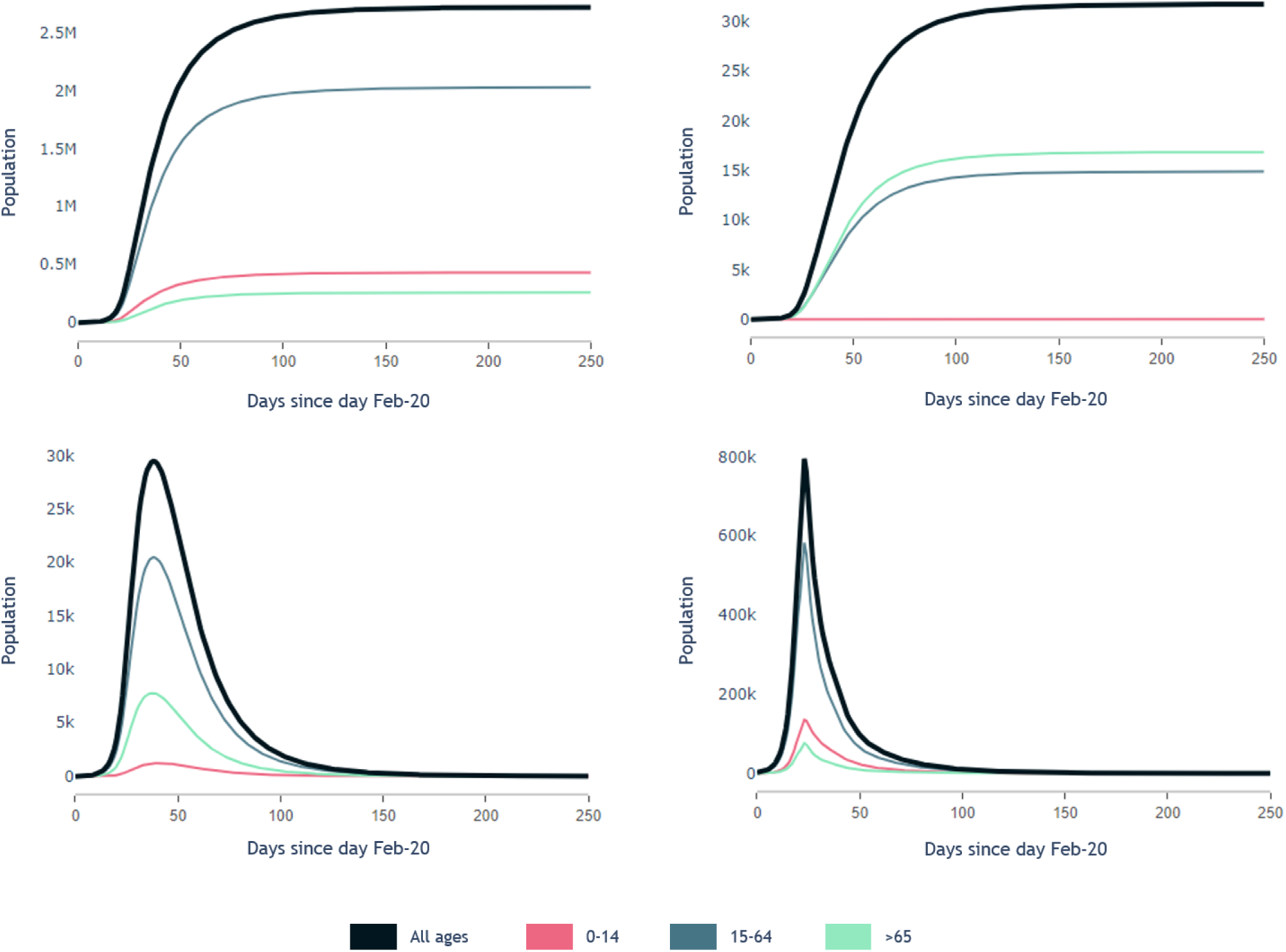
Mobility restricted up to 30% of movements and contained during the following 250 days. From left to right, up to down, first graph shows accumulated number of people who recovered after contracting COVID-19, second graph shows accumulated number of people who deceased by COVID-19, third graph shows instantaneous number of people making use of health services and fourth graph shows instantaneous asymptomatic people.

## Data Availability

People's mobility data was extracted from the Spanish National Statistics institute.
Covid-19 cases in Spain were extracted from the Spanish National Healthcare Ministery.

https://www.mscbs.gob.es/profesionales/saludPublica/ccayes/alertasActual/nCov/situacionActual.htm

https://www.ine.es/

## Acknowledgments

We thank the Universidad Politecnica de Madrid (UPM) for supporting this study. We also thank the Instituto Tecnológico de Aragón (ITAINNOVA) for the useful discussions.

